# The potential impact of reduced international donor funding on the household economic burden of tuberculosis in low- and middle-income countries

**DOI:** 10.1101/2025.08.09.25333356

**Authors:** Allison Portnoy, Rebecca A. Clark, Mark Jit, C. Finn McQuaid, Alexandra S. Richards, Roel Bakker, Tom Sumner, Tomos O. Prŷs-Jones, Rein M.G.J. Houben, Richard G. White, Katherine C. Horton, Nicolas A. Menzies

## Abstract

**Background:** Recent shifts in the global health funding landscape—most notably the dismantling of the United States Agency for International Development (USAID) and possible reduced contributions to the Global Fund to Fight AIDS, TB and Malaria (Global Fund)— threaten essential tuberculosis (TB) services in low- and middle-income countries (LMICs). We quantified the potential impact on the household economic burden of TB.

**Methods:** We used linked epidemiological and economic models, calibrated to 79 LMICs, to estimate future TB patient costs under four scenarios: continuation of 2024 funding levels (baseline), termination of USAID, termination of USAID plus announced reductions in Global Fund contributions, and full elimination of external funding for TB. Outcomes included total TB-attributable household costs and numbers of households experiencing catastrophic costs (disease-related costs >20% of annual income).

**Findings:** USAID termination was projected to produce US$7.5 (95% uncertainty interval: $6.1–8.9) billion in additional patient-incurred costs and 3.9 (3.1–4.6) million additional households experiencing catastrophic costs over 2025–2050. The worst-case scenario (elimination of all external funding) resulted in $79.7 ($60.0–99.2) billion in additional patient-incurred costs and 40.5 (30.9–50.7) million additional households experiencing catastrophic costs—a 32% increase over baseline. Impacts were greatest for poorer households, with over 50% of additional catastrophic costs occurring in the poorest 20% of households.

**Interpretation:** Abrupt reductions in international donor funding for TB may reverse recent progress toward financial risk protection and health equity in LMICs. Strategies to reduce the disruption caused by funding cuts and protect vulnerable populations are urgently needed.

**Research in context:** *Evidence before this study:* Several prior studies have examined the potential impact of cuts in international health funding from the United States of America. We searched PubMed and medRxiv for studies quantifying the effects of reductions in international donor funding on the economic burden of tuberculosis, published between January 1 and August 7, 2025, using search terms related to funding (“funding”, “donor”, “aid”, “assistance”), tuberculosis (“tuberculosis”, “TB”), patients or households (patient*, household*), and economic burden (cost*, econ*). The identified studies described a range of potential health consequences that could result from funding cuts. To our knowledge, no studies have considered the impact of funding cuts on the household economic burden of disease.

*Added value of this study:* Our modelling suggested that termination of United States Agency for International Development (USAID) funding could lead to US$7.5 (95% uncertainty interval: $6.1–8.9) billion in additional patient-incurred costs and 3.9 (3.1–4.6) million additional households experiencing catastrophic costs over 2025–2050. Further reductions in funding to the Global Fund to Fight AIDS, TB and Malaria (Global Fund) in line with current announcements from donor countries could lead to a further $21.2 ($16.6–25.6) billion in patient-incurred costs and 10.7 (8.4–13.0) million households experiencing catastrophic costs. If all external TB funding were terminated, a projected $72.2 ($53.9–90.4) billion in patient-incurred costs could accrue and 36.6 (27.7–46.1) million households could experience catastrophic costs, compared with the impact of the funding cuts to USAID alone.

*Implications of all the available evidence:* Disruptions to TB services resulting from reductions in international donor funding could result in increased tuberculosis-associated morbidity and mortality, which in turn could result in increased economic burden on resource-constrained households in the world’s poorest countries. Strategies to reduce the disruption caused by funding cuts and protect vulnerable populations are urgently needed.

## Introduction

Tuberculosis (TB) is the world’s leading cause of death from an infectious agent, responsible for an estimated 1.25 million deaths in 2023.^1^ Despite slow declines in TB incidence since 2000, global rates have been increasing since 2020—approximately 10.8 million individuals are estimated to have developed TB in 2023, up from 10.1 million in 2020.^1^ The impact of this morbidity and mortality falls disproportionately on low- and middle-income countries (LMIC), with 30 LMICs with the highest TB burden accounting for 87% of overall incidence.^1,2^

The costs incurred by individuals who develop TB place a high financial burden on TB-affected households in LMIC settings. These costs arise from the out-of-pocket expenses (such as transport and accommodation) required to participate in TB treatment, as well as the income losses incurred by individuals who are unable to work. Due to these two factors, more than 50% of TB-affected households face total costs during the TB episode that are greater than 20% of annual household income, and therefore categorized as catastrophic costs by the WHO.^1^ These impacts are greater for TB-affected households in the poorest income quintile, with 75% of these households incurring catastrophic costs.^3^ Reducing the costs faced by patients and their households is a high-level objective of the World Health Organization’s (WHO) End TB Strategy, yet progress to eliminate catastrophic costs due to TB has been slow.^4^

International donor funding has played an important role historically in supporting access to TB prevention and treatment services, which contribute to reducing household economic burden in resource-limited settings. Until early 2025, the primary international funders of the global TB response were the United States Agency for International Development (USAID), which provided 19% of international donor funding reported by national TB programs (NTPs) in 2023, and the Global Fund to Fight AIDS, TB and Malaria (Global Fund), which provided 76% of international donor funding—38% contributed by the United States of America—in the same year.^1,5^ The financial support of these agencies, led by the largest single country donor (the United States of America), enabled progress towards closing the diagnostic and treatment gaps faced by people with TB and their households, and mitigating the high financial costs faced by TB patients.

Recent progress in combatting TB is now threatened by changes to the international donor funding landscape. In early 2025, the United States abruptly dismantled USAID, cancelling bilateral health aid provided by the agency.^6^ The ramifications of this loss of funding have already interrupted TB prevention, diagnostic, and treatment services.^7,8^ Budget proposals within the U.S. government in July 2025 proposed substantially reduced support for international health programs, including the Global Fund. In addition to the United States, other donor countries have announced reductions in contributions to bilateral aid programs and multinational organizations.^6^

In this study, we examined how changes in donor funding could impact the economic burden of TB on affected households in LMICs. To do so we extended an earlier study that projected the potential health impact of funding reductions,^9^ to quantify the household economic consequences of TB, the number of TB-affected households experiencing catastrophic costs, and the distribution of these outcomes across income quintiles in each country over the period 2025– 2050. We conducted this analysis for 79 LMICs—representing 91% of TB incidence in LMICs.

## Methods

### Scenarios

We examined three scenarios describing different patterns of future funding for the TB response in the 79 LMICs included in the analysis.^9^ First, we simulated a scenario representing the impact of the termination of USAID funding to NTPs beginning in 2025. Second, we simulated a scenario representing the impact of both the termination of USAID funding and announced reductions in funding to the Global Fund, equivalent to a 46% reduction in total Global Fund contributions. Finally, we examined a scenario estimating the impact of complete termination of both USAID and Global Fund funding, as well as termination of external TB funding from other non-government and private organizations. Each scenario was compared with a baseline scenario assuming TB funding and programmatic activities continue at 2024 levels.

We assumed all funding cuts would be introduced in 2025, sustained into the future, and not replaced by other funding sources, and that unless specified in a scenario other funding sources (domestic, other international) would continue at 2024 levels (additional details reported in Clark, et al.).^9^ We assumed the impact of reduced funding for TB programs could be represented by reductions in access to TB diagnosis and treatment. We operationalized this by assuming there would be a reduction in the TB treatment initiation rate (for individuals with untreated TB disease) that is proportional to the budget reduction represented by a given scenario. These budget components were estimated from expenditure data reported by countries to the WHO, which has monitored funding for TB programs since 2022.^1^ For each country, WHO data were used to calculate the proportion of total expected funding from all sources from USAID, from the Global Fund, and from other external donors in 2023.

### Mathematical model

To undertake the analysis we adapted a system of linked epidemiological and economic models originally developed to examine the implications of global TB vaccine introduction.^10–12^ Models were calibrated to demographic, epidemiological and health service data for 79 LMICs.^9^

### Costs incurred by TB-affected households

For each modelled country, we disaggregated the TB burden projected under each scenario by income quintile, based on survey data describing differences in the relative prevalence of TB by household income.^12^ To calculate the costs incurred by TB patients and their households we multiplied the simulated number of TB cases (by country, income quintile, scenario, and year) by the average patient cost per TB episode. Estimates of the patient costs per TB episode, stratified by country and household income quintile, were derived from a published meta-regression analysis of 22 nationally representative TB patient cost surveys.^3^ These estimates included direct medical costs (medical products and services), direct non-medical costs (travel, accommodation, food, and nutritional supplements) and indirect costs (income losses) incurred by TB patients. For each country and income quintile, we assumed that the per-patient costs of TB (in 2021 constant dollars) would not change in future years. For the base-case analysis, we assumed that individuals with TB disease who do not receive appropriate treatment (i.e., received ineffective care, or not treated at all) experience the same total per episode costs as those who received appropriate treatment. We examined alternative assumptions in sensitivity analyses.

### Catastrophic costs

As defined in the WHO End TB Strategy targets, we calculated catastrophic costs as instances where the patient costs of TB disease—the sum of direct medical costs, direct non-medical costs and indirect costs—exceeded 20% of total annual income for the TB-affected household.^13–16^ For each country and income quintile, we assessed the number of TB-affected households experiencing catastrophic costs under each scenario, multiplying the probability of catastrophic costs per TB episode by the simulated number of TB cases by country, income quintile, scenario, and year. Estimates of the probability of catastrophic costs per TB episode (stratified by country and income quintile) were derived from the meta-analysis of TB patient cost surveys used for patient cost estimates.^3^ For each country and income quintile, we assumed that the probability of catastrophic costs for TB patients would not change in future years.

### Distribution of benefits across countries and income strata

We undertook additional analyses to describe how the number of households experiencing catastrophic costs was distributed across the collective income gradient of the modelled countries. First, we ordered all country income quintiles (79 countries × 5 quintiles = 395 unique groups) by average per capita gross domestic product (GDP) in 2021 purchasing power parity (PPP)-adjusted dollars. To do so, we obtained estimates of per capita PPP GDP and the fraction of total income held by each country income quintile, imputing missing values according to WHO region and income level group averages (e.g., low-income countries in the African region).^17,18^ We multiplied these two terms and divided by the population fraction in each quintile (0.2) to obtain the average per capita PPP GDP for each quintile. We ranked all quintiles by average per capita PPP GDP and calculated the distribution of each study outcome across these quintiles. We summarized results graphically as well as via the Concentration Index, which quantifies the relative concentration of a given outcome in high-income or low-income groups. For a cumulative distribution representing incidence of an outcome in percentiles of a population ordered from poorest to richest (the ‘Concentration Curve’), the Concentration Index is defined as two times the area between the Concentration Curve and the line of equality (the 45° line, representing an equal distribution of the outcome across income groups). The index is defined in [−1, 1], with more positive (negative) values indicating greater concentration of the outcome in higher (lower)-income groups.^19^

### Sensitivity analysis

We propagated uncertainty in analytic inputs using a second-order Monte Carlo simulation, which generated 200 estimates for each outcome. We used this distribution of estimates to generate equal-tailed 95% uncertainty intervals for each study outcome. We also examined the robustness of results to alternative analytic assumptions. First, as there is substantial uncertainty around the costs incurred by patients who do not receive appropriate TB treatment, we re-estimated results under alternative scenarios that assumed costs for this group were 50% lower and higher, respectively, compared with appropriately treated individuals (vs. the main analysis which assumed treated and untreated patients bore the same costs). Second, we examined alternative thresholds for defining catastrophic costs as 10% and 25% of annual household income (vs 20% in the main analysis).

## Results

### Overall potential impact of funding cuts

Compared with the baseline scenario, the termination of USAID funding in the 79 modelled countries was projected to result in $7.5 (95% uncertainty interval: $6.1–8.9) billion in additional costs borne by TB-affected households, including $1.2 ($0.8–1.5) billion in direct medical costs, $2.8 ($2.1–3.4) billion in direct non-medical costs, and $3.6 ($2.8–4.6) billion in indirect costs over the 2025–2050 study period (Table 1). For this scenario, we estimated 3.9 (3.1–4.6) million additional households would experience catastrophic costs, as compared to the baseline. Reductions in funding to the Global Fund in addition to the termination of USAID could result in approximately $28.7 ($22.7–34.4) billion in increased costs to TB-affected households, and an additional 14.6 (11.6–17.6) million households experiencing catastrophic costs due to TB (a 12% increase compared to baseline).

**Table 1.** Costs borne by TB-affected households and number of households with catastrophic costs.

| Scenario | Increased patient direct medical costs (billions) | Increased patient direct non-medical costs (billions) | Increased patient indirect costs (billions) | Increased patient total costs (billions) | Increased cases of catastrophic costs (millions) |
| --- | --- | --- | --- | --- | --- |
| Termination of USAID | 1.2 (0.8–1.5)<br>*2.3% (1.7–3.0%)* | 2.8 (2.1–3.4)<br>*2.3% (1.7–2.9%)* | 3.6 (2.8–4.6)<br>*2.7% (2.2–3.2%)* | 7.5 (6.1–8.9)<br>*2.5% (2.0–2.9%)* | 3.9 (3.2–4.6)<br>*3.1% (2.5–3.6%)* |
| Termination of USAID and reduced Global Fund contributions | 4.6 (3.2–6.1)<br>*9.2% (6.7–11.9%)* | 10.8 (7.8–13.6)<br>*8.8% (6.3–11.3%)* | 13.2 (10.0–16.8)<br>*9.9% (8.0–12.0%)* | 28.7 (22.7–34.4)<br>*9.4% (7.2–11.5%)* | 14.6 (11.6–17.6)<br>*11.5% (9.0–14.1%)* |
| Termination of all external TB funding | 13.2 (8.7–18.1)<br>*26.3% (19.4–34.4%)* | 30.1 (20.7–39.5)<br>*24.6% (16.5–33.4%)* | 36.4 (26.4–47.3)<br>*27.3% (20.8–35.0%)* | 79.7 (60.0–99.2)<br>*26.0% (19.2–33.8%)* | 40.5 (30.9–50.7)<br>*31.9% (24.2–41.3%)* |

Note: Estimates include 79 low- and middle-income countries analyzed compared to the baseline scenario. Values in italicized text represent percentage changes compared to the baseline scenario. Values in parentheses represent equal-tailed 95% credible intervals. Total costs included patient direct medical, direct non-medical, and indirect costs (all undiscounted) over 2025–2050 in 2021 US dollars. Catastrophic costs are defined as instances where the total patient costs incurred during an episode of TB disease exceeded 20% of total annual household income. GF = Global Fund to Fight AIDS, TB and Malaria; TB=tuberculosis; USAID = United States Agency of International Development.

If all external TB funding were terminated, we projected an increase of $79.7 ($60.0–99.2) billion in total patient costs, including $13.2 ($8.7–18.1) billion in direct medical costs, $30.1 ($20.7–39.5) billion in direct non-medical costs, and $36.4 ($26.4–47.3) billion in indirect costs. This would result in an estimated 40.5 (30.9–50.7) million additional cases of households experiencing catastrophic costs, an increase of nearly 32% compared to what was projected under the baseline.

### Distribution of TB patient costs within each country

Across all modelled countries and scenarios, the wealthiest two income quintiles accounted for approximately 51% of increased total patient costs resulting from cuts to international donor funding (Concentration Index 0.14 for each scenario), with greater costs per episode of TB care incurred in these groups outweighing the greater burden of TB cases in poorer quintiles.

The largest absolute increases in the proportion of households facing catastrophic costs were in lower income quintiles within each country. Under each scenario, nearly 60% of cases of catastrophic costs were experienced by the poorest two quintiles (Concentration Index −0.24 for each scenario). Figure 1 shows the relative magnitude of cases of catastrophic costs averted across income quintiles by analytic scenario. The percentage of TB-affected households experiencing catastrophic costs by country GDP per capita and by income quintile is presented in appendix Figure S1.

**Figure 1.**
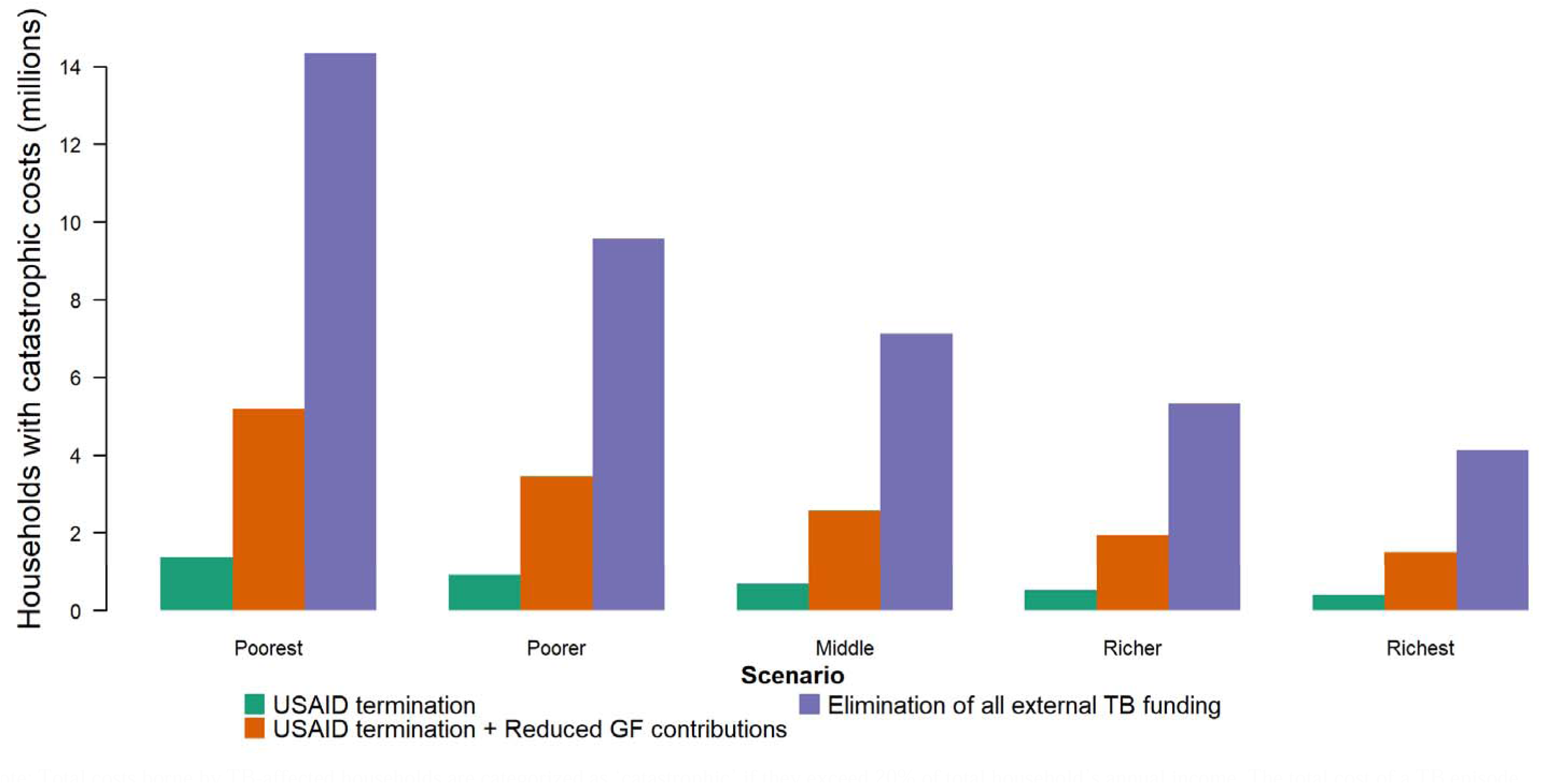
Number of increased tuberculosis (TB)-affected households experiencing catastrophic costs by within-country income quintile, compared to no changes in donor funding. Note: Total costs borne by TB-affected households are categorized as ‘catastrophic’ if they exceed 20% of total household’s annual income. The total cost of a TB episode included patient direct medical, direct non-medical and indirect costs over 2025–2050. GF = The Global Fund to Fight AIDS, TB and Malaria; USAID = United States Agency of International Development.

### Distribution of outcomes across countries and income strata

Figure 2 shows the distribution of TB-affected households experiencing catastrophic costs by household income across the combined population of the 79 modelled LMICs over the 2025– 2050 period for the three scenarios, each compared to a counterfactual scenario without funding losses. In all scenarios, expected increases to the number of households experiencing catastrophic costs were concentrated in poorer households (poorest quintile shaded in red), with 68% (Concentration Index −0.61) and 59% of the total cases of catastrophic costs in the poorest 20% of households (Concentration Index −0.53), respectively.

**Figure 2.**
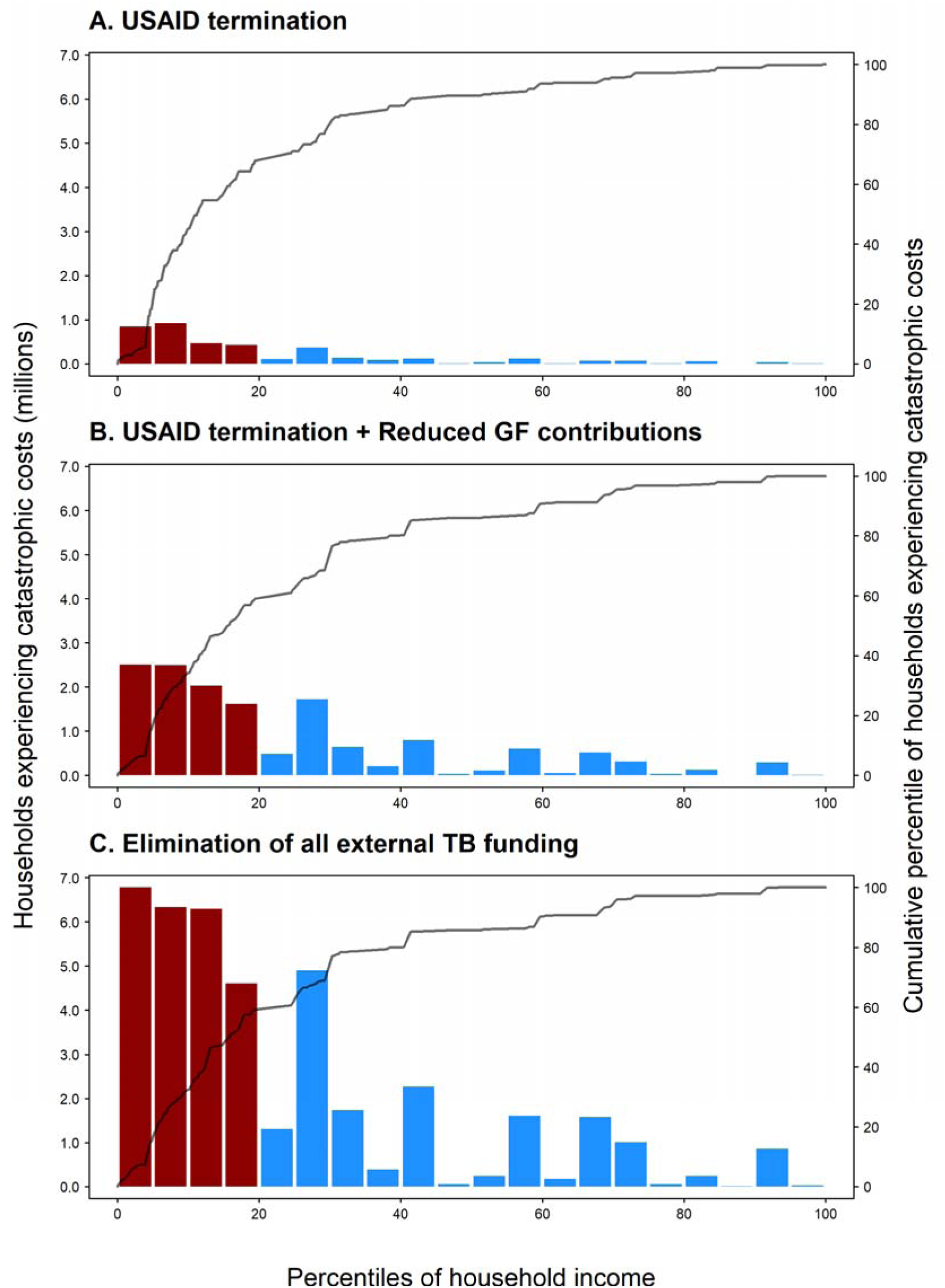
Distribution of the number of TB-affected households experiencing catastrophic costs over 2025–2050 under alternative scenarios for possible reductions in external TB funding, across all modelled strata, ordered by household income. Note: GDP: gross domestic product; GF: the Global Fund to Fight AIDS, TB and Malaria; TB: tuberculosis; USAID: the United States Agency of International Development. Bars defined by left-hand side y-axis; lines defined by right-hand side y-axis. Ordering of population by household income based on average 2021 per capita GDP in purchasing power parity (PPP) dollars, for each modelled stratum (395 total strata). Bars shaded red indicate the poorest 20% of modelled population by PPP GDP per capita.

### Sensitivity analyses

Assuming that individuals with TB who do not receive appropriate treatment experience 50% lower costs compared with treated individuals reduced the number of households with catastrophic costs for each scenario (Table S1).

Assuming that individuals with TB who do not receive appropriate treatment experience 50% higher costs compared with treated individuals increased the number of households with catastrophic costs for each scenario (Table S2).

Defining catastrophic costs as instances where patient costs exceed 10% of annual household income (vs 20% in the main analysis), resulted in approximately 39% more cases of catastrophic costs, as compared to the main analysis (Table S3). Defining catastrophic costs as instances where patient costs exceed 25% of annual household income resulted in approximately 14% fewer cases of catastrophic costs compared to the main analysis.

## Discussion

This study quantifies the potential impact of reduced health aid by the United States and other countries on the costs faced by TB-affected households in low- and middle-income countries. In a worst-case scenario of the full termination of external TB funding, the results of this analysis suggest that TB-affected households could face $79.7 ($60.0–99.2) billion in additional direct medical, non-medical, and indirect costs for TB care, resulting in an estimated 40.5 (30.9–50.7) million additional households experiencing catastrophic costs.

Sharp reductions in international donor funding pose significant threats to the continuity and quality of essential health services in LMICs. While external funding has played a transformative role in scaling up TB prevention and treatment, the abrupt change in the funding landscape since the beginning of 2025 has already led to service disruptions.^6–8^ The withdrawal of donor support could erode healthcare access, particularly for the poorest households that experience the greatest burden of TB.^12^ The timeline to achieve the End TB Strategy target to eliminate household catastrophic costs could also be substantially lengthened.^4^ This study demonstrates that in addition to the health losses that could result from funding reductions,^9^ the projected economic burden that households will face given a sharp withdrawal of funding without alternative financing mechanisms could be catastrophic.

For the sustainability of national TB programs, blended financing models that combine public, private, and donor resources may offer interim solutions, particularly in countries with constrained fiscal space, as countries work towards strengthening domestic resource mobilization. Additionally, there may be opportunities to integrate aspects of national TB programs into primary health care in order to promote efficiency and enhance system resilience.

This analysis has several limitations. First, there is substantial uncertainty around the costs faced by individuals with TB who are not able to access treatment. While these individuals will incur substantial costs associated with care-seeking that does not end in appropriate treatment, as well as income losses from untreated TB morbidity, these is little empirical evidence quantifying the magnitude of these costs. In the main analysis we assumed that untreated individuals experienced the same costs as treated individuals, based on limited evidence highlighting costs faced by these individuals.^20^ We then examined alternative assumptions for the costs faced by untreated individuals as well as alternative thresholds for defining catastrophic costs as a percentage of total annual household income in sensitivity analysis. Second, we assumed that the current estimated costs of each episode for TB patients would not change over the 2025–2050 period (in constant US dollars). If the patient costs per TB episode were to decline, or real incomes rise, this would reduce the number of households experiencing catastrophic costs due to TB. Third, we assumed a standard reduction in treatment initiation for all TB patients in the underlying epidemiological modelling informing this analysis, but the impact on treatment initiation could differ based on patient characteristics, including household income. Fourth, we did not consider secondary effects of international donor funding cuts to other sectors on NTPs. The long-term effects of funding cuts (e.g., to nutrition, HIV programs) could further impact the number of households with TB and/or the number of households at risk of catastrophic costs due to TB-associated care. Finally, the funding scenarios we examined were deliberately simplified to reveal the potential impact of current and possible funding cuts. It is unlikely that the trajectory of externally health funding for the modelled countries will exactly match one of the scenarios examined in this analysis, but these scenarios represent extremes describing the possible range of consequences. The findings of this study can contribute to the ongoing policy debates surrounding the risks and opportunities inherent in the shift from externally financed to domestically supported health systems. As described by Pai et al. (2024),^21^ donors should support transition processes through flexible, phased approaches that allow for country-specific adaptation and a fair share of overhead costs. Global health agencies can play a facilitative role by providing technical assistance and capacity strengthening for partner institutions in LMICs. For LMIC governments, it is essential to invest in institutional capacity and enhance coordination across sectors and levels of government. Ultimately, a shared commitment to equity, sustainability, and health system strengthening must not only guide the transition toward more domestically anchored and resilient health systems, but also ensure due consideration to protect vulnerable populations in the world’s poorest countries in the face of reduced global health resources expected in coming years.

## Data Availability

All data produced in the present study are available upon reasonable request to the authors.

---

**Table S1.** Costs borne by TB-affected households and number of households with catastrophic costs (in millions), assuming costs for untreated cases are 0.5x treated cases.

| Scenario | Increased patient direct medical costs | Increased patient direct non-medical costs | Increased patient indirect costs | Increased patient total costs | Increased cases of catastrophic costs |
| --- | --- | --- | --- | --- | --- |
| Termination of USAID | 0.5 (0.3–0.8)<br>*1.2% (0.7–1.7%)* | 1.2 (0.6–1.7)<br>*1.1% (0.6–1.7%)* | 1.7 (1.0–2.5)<br>*1.4% (0.8–2.0%)* | 3.4 (2.1–4.6)<br>*1.2% (0.7–1.7%)* | 2.3 (1.9–2.7)<br>*3.2% (2.6–3.8%)* |
| Termination of USAID and reduced Global Fund contributions | 1.9 (1.1–2.9)<br>*4.3% (2.4–6.4%)* | 4.5 (2.1–6.7)<br>*4.2% (2.1–6.7%)* | 5.7 (2.9–8.6)<br>*4.7% (2.5–6.9%)* | 12.1 (6.2–17.6)<br>*4.4% (2.3–6.6%)* | 8.7 (7.0–10.3)<br>*11.9% (9.4–14.3%)* |
| Termination of all external TB funding | 3.7 (1.4–6.5)<br>*8.3% (3.2–14.4%)* | 8.8 (2.6–15.5)<br>*8.2% (2.8–14.7%)* | 10.1 (3.6–17.3)<br>*8.3% (3.2–14.6%)* | 22.5 (7.4–37.7)<br>*8.3% (2.7–14.4%)* | 24.1 (18.6–30.1)<br>*33.1% (25.2–42.4%)* |

Note: Estimates include 79 low- and middle-income countries analyzed compared to the baseline scenario. Values in italicized text represent percentage changes compared to the baseline scenario. Values in parentheses represent equal-tailed 95% credible intervals. Total costs included patient direct medical, direct non-medical, and indirect costs (all undiscounted) over 2025–2050 in 2021 USD. Catastrophic costs are defined as instances where the total patient costs incurred during an episode of TB disease exceeded 20% of total annual household income. GF = Global Fund to Fight AIDS, TB and Malaria; TB=tuberculosis; USAID = United States Agency of International Development.

**Table S2.** Costs borne by TB-affected households and number of households with catastrophic costs (in millions), assuming costs for untreated cases are 1.5x treated cases.

| Scenario | Increased patient direct medical costs | Increased patient direct non-medical costs | Increased patient indirect costs | Increased patient total costs | Increased cases of catastrophic costs |
| --- | --- | --- | --- | --- | --- |
| Termination of USAID | 1.8 (1.3–2.4)<br>*3.2% (2.6–4.0%)* | 4.3 (3.3–5.2)<br>*3.1% (2.4–3.8%)* | 5.5 (4.4–6.8)<br>*3.8% (3.2–4.4%)* | 11.7 (10.1–13.4)<br>*3.4% (2.9–4.0%)* | 4.8 (3.9–5.6)<br>*3.0% (2.4–3.5%)* |
| Termination of USAID and reduced Global Fund contributions | 7.4 (5.3–9.5)<br>*13.1% (10.1–16.2%)* | 17.1 (13.5–20.7)<br>*12.5% (9.4–15.3%)* | 20.7 (16.5–25.2)<br>*14.3% (12.0–16.5%)* | 45.2 (38.2–52.1)<br>*13.3% (11.1–15.6%)* | 18.1 (14.3–21.8)<br>*11.3% (8.8–13.9%)* |
| Termination of all external TB funding | 22.8 (16.1–29.7)<br>*40.5% (30.8–50.7%)* | 51.4 (38.4–64.6)<br>*37.4% (27–47.9%)* | 62.7 (48.1–78.5)<br>*43.3% (35.1–52.4%)* | 136.8 (112.3–161.2)<br>*40.4% (32.1–49.4%)* | 50.1 (38.0–63.0)<br>*31.2% (23.7–40.6%)* |

Note: Estimates include 79 low- and middle-income countries analyzed compared to the baseline scenario. Values in italicized text represent percentage changes compared to the baseline scenario. Values in parentheses represent equal-tailed 95% credible intervals. Total costs included patient direct medical, direct non-medical, and indirect costs (all undiscounted) over 2025–2050 in 2021 USD. Catastrophic costs are defined as instances where the total patient costs incurred during an episode of TB disease exceeded 20% of total annual household income. GF = Global Fund to Fight AIDS, TB and Malaria; TB=tuberculosis; USAID = United States Agency of International Development.

**Table S3.**
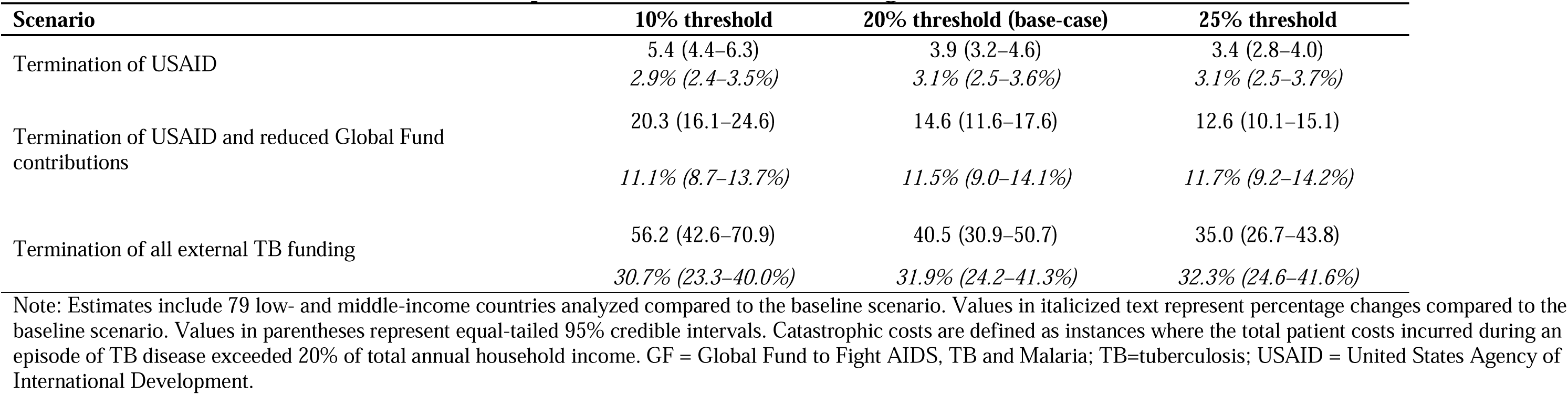
Number of households with catastrophic costs (in millions), assuming thresholds of 10%, 20% (base-case), and 25%.

**Figure S1.**
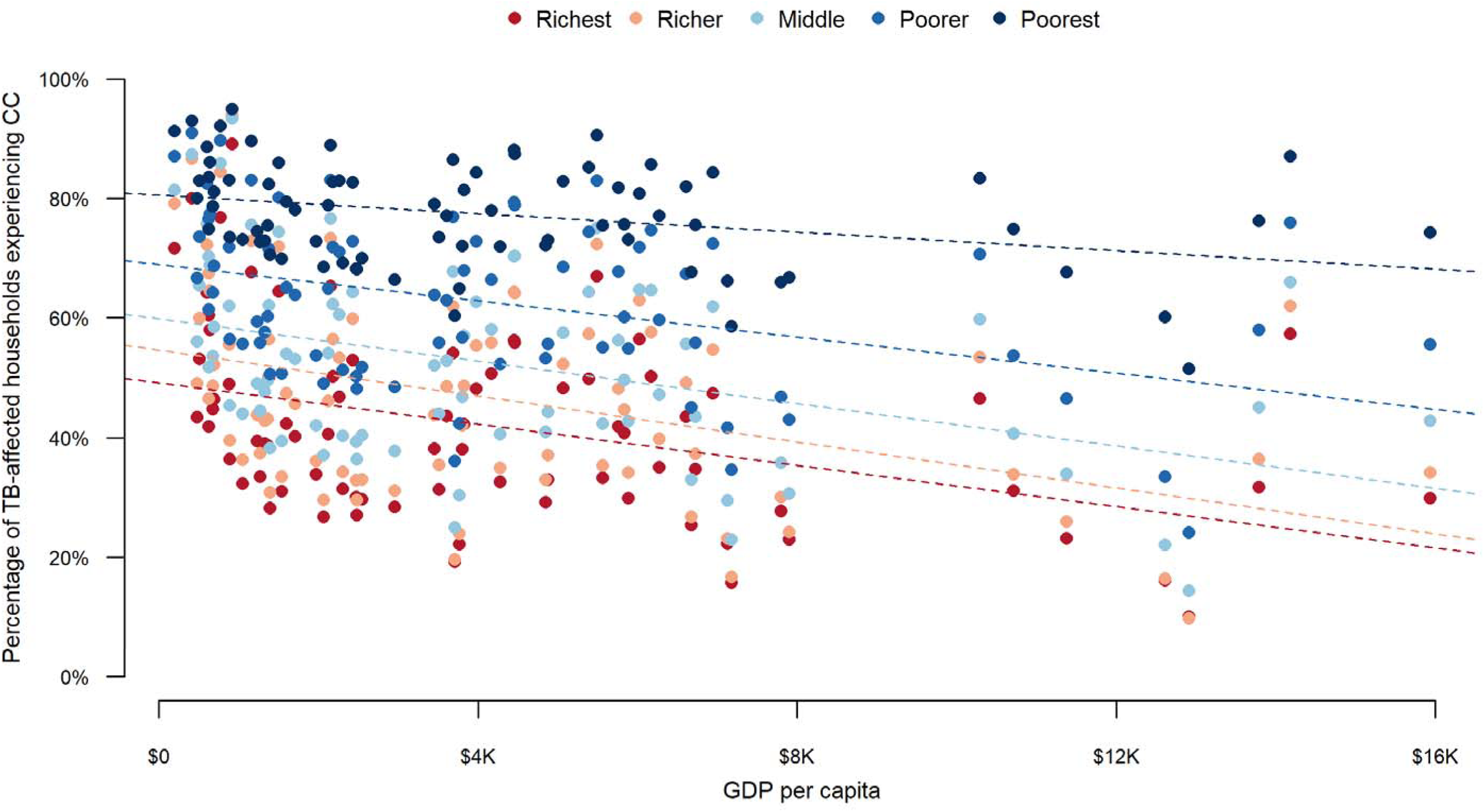
Percentage of tuberculosis (TB)-affected households experiencing catastrophic costs (CC) by gross domestic product (GDP) per capita and by income quintile in 79 low- and middle-income countries. Note: Dotted lines represent trends for the percentage of TB-affected households experiencing catastrophic costs by GDP per capita for each income quintile. Colors represent income quintiles for each analyzed country.

