## Supplementary Appendix for "The potential impact of reduced international donor funding on the household economic burden of tuberculosis in low- and middle-income countries"

### Table S1. Costs borne by TB-affected households and number of households with catastrophic costs (in millions), assuming costs for untreated cases are 0.5x treated cases.

| **Scenario** | **Increased patient direct medical costs** | **Increased patient direct non–medical costs** | **Increased patient indirect costs** | **Increased patient total costs** | **Increased cases of catastrophic costs** |
| --- | --- | --- | --- | --- | --- |
| Termination of USAID | 0.5 (0.3–0.8) | 1.2 (0.6–1.7) | 1.7 (1.0–2.5) | 3.4 (2.1–4.6) | 2.3 (1.9–2.7) |
|  | *1.2% (0.7–1.7%)* | *1.1% (0.6–1.7%)* | *1.4% (0.8–2.0%)* | *1.2% (0.7–1.7%)* | *3.2% (2.6–3.8%)* |
| Termination of USAID and reduced Global Fund contributions | 1.9 (1.1–2.9) | 4.5 (2.1–6.7) | 5.7 (2.9–8.6) | 12.1 (6.2–17.6) | 8.7 (7.0–10.3) |
|  | *4.3% (2.4–6.4%)* | *4.2% (2.1–6.7%)* | *4.7% (2.5–6.9%)* | *4.4% (2.3–6.6%)* | *11.9% (9.4–14.3%)* |
| Termination of all external TB funding | 3.7 (1.4–6.5) | 8.8 (2.6–15.5) | 10.1 (3.6–17.3) | 22.5 (7.4–37.7) | 24.1 (18.6–30.1) |
|  | *8.3% (3.2–14.4%)* | *8.2% (2.8–14.7%)* | *8.3% (3.2–14.6%)* | *8.3% (2.7–14.4%)* | *33.1% (25.2–42.4%)* |

Note: Estimates include 79 low- and middle-income countries analyzed compared to the baseline scenario. Values in italicized text represent percentage changes compared to the baseline scenario. Values in parentheses represent equal-tailed 95% credible intervals. Total costs included patient direct medical, direct non-medical, and indirect costs (all undiscounted) over 2025–2050 in 2021 USD. Catastrophic costs are defined as instances where the total patient costs incurred during an episode of TB disease exceeded 20% of total annual household income. GF = Global Fund to Fight AIDS, TB and Malaria; TB=tuberculosis; USAID = United States Agency of International Development.

### Table S2. Costs borne by TB-affected households and number of households with catastrophic costs (in millions), assuming costs for untreated cases are 1.5x treated cases.

| **Scenario** | **Increased patient direct medical costs** | **Increased patient direct non–medical costs** | **Increased patient indirect costs** | **Increased patient total costs** | **Increased cases of catastrophic costs** |
| --- | --- | --- | --- | --- | --- |
| Termination of USAID | 1.8 (1.3–2.4) | 4.3 (3.3–5.2) | 5.5 (4.4–6.8) | 11.7 (10.1–13.4) | 4.8 (3.9–5.6) |
|  | *3.2% (2.6–4.0%)* | *3.1% (2.4–3.8%)* | *3.8% (3.2–4.4%)* | *3.4% (2.9–4.0%)* | *3.0% (2.4–3.5%)* |
| Termination of USAID and reduced Global Fund contributions | 7.4 (5.3–9.5) | 17.1 (13.5–20.7) | 20.7 (16.5–25.2) | 45.2 (38.2–52.1) | 18.1 (14.3–21.8) |
|  | *13.1% (10.1–16.2%)* | *12.5% (9.4–15.3%)* | *14.3% (12.0–16.5%)* | *13.3% (11.1–15.6%)* | *11.3% (8.8–13.9%)* |
| Termination of all external TB funding | 22.8 (16.1–29.7) | 51.4 (38.4–64.6) | 62.7 (48.1–78.5) | 136.8 (112.3–161.2) | 50.1 (38.0–63.0) |
|  | *40.5% (30.8–50.7%)* | *37.4% (27–47.9%)* | *43.3% (35.1–52.4%)* | *40.4% (32.1–49.4%)* | *31.2% (23.7–40.6%)* |

Note: Estimates include 79 low- and middle-income countries analyzed compared to the baseline scenario. Values in italicized text represent percentage changes compared to the baseline scenario. Values in parentheses represent equal-tailed 95% credible intervals. Total costs included patient direct medical, direct non-medical, and indirect costs (all undiscounted) over 2025–2050 in 2021 USD. Catastrophic costs are defined as instances where the total patient costs incurred during an episode of TB disease exceeded 20% of total annual household income. GF = Global Fund to Fight AIDS, TB and Malaria; TB=tuberculosis; USAID = United States Agency of International Development.

### Table S3. Number of households with catastrophic costs (in millions), assuming thresholds of 10%, 20% (base-case), and 25%.

| **Scenario** | **10% threshold** | **20% threshold (base-case)** | **25% threshold** |
| --- | --- | --- | --- |
| Termination of USAID | 5.4 (4.4–6.3) | 3.9 (3.2–4.6) | 3.4 (2.8–4.0) |
|  | *2.9% (2.4–3.5%)* | *3.1% (2.5–3.6%)* | *3.1% (2.5–3.7%)* |
| Termination of USAID and reduced Global Fund contributions | 20.3 (16.1–24.6) | 14.6 (11.6–17.6) | 12.6 (10.1–15.1) |
|  | *11.1% (8.7–13.7%)* | *11.5% (9.0–14.1%)* | *11.7% (9.2–14.2%)* |
| Termination of all external TB funding | 56.2 (42.6–70.9) | 40.5 (30.9–50.7) | 35.0 (26.7–43.8) |
|  | *30.7% (23.3–40.0%)* | *31.9% (24.2–41.3%)* | *32.3% (24.6–41.6%)* |

Note: Estimates include 79 low- and middle-income countries analyzed compared to the baseline scenario. Values in italicized text represent percentage changes compared to the baseline scenario. Values in parentheses represent equal-tailed 95% credible intervals. Catastrophic costs are defined as instances where the total patient costs incurred during an episode of TB disease exceeded 20% of total annual household income. GF = Global Fund to Fight AIDS, TB and Malaria; TB=tuberculosis; USAID = United States Agency of International Development.

### Figure S1. Percentage of tuberculosis (TB)-affected households experiencing catastrophic costs (CC) by gross domestic product (GDP) per capita and by income quintile in 79 low- and middle-income countries.

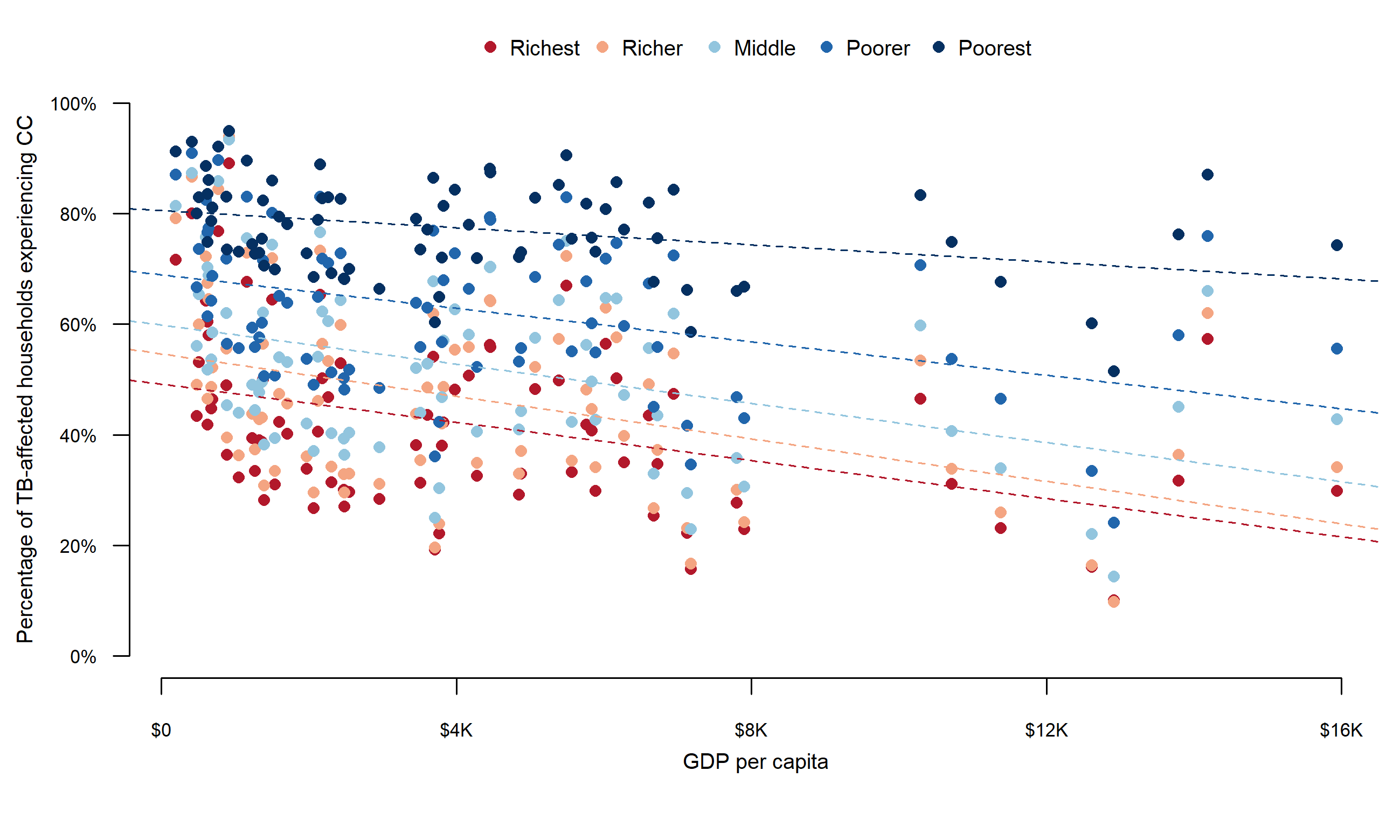

**Note: Dotted lines represent trends for the percentage of TB-affected households experiencing catastrophic costs by GDP per capita for each income quintile. Colors represent income quintiles for each analyzed country.**
